# Performance of SARS-CoV-2 rapid antigen test compared with real-time RT-PCR in asymptomatic individuals

**DOI:** 10.1101/2021.02.12.21251643

**Authors:** Mónica Peña, Manuel Ampuero, Carlos Garcés, Aldo Gaggero, Patricia García, María Soledad Velasquez, Ricardo Luza, Pía Alvarez, Fabio Paredes, Johanna Acevedo, Mauricio J. Farfán, Sandra Solari, Ricardo Soto-Rifo, y Fernando Valiente-Echeverría

## Abstract

Screening, testing and contact tracing plays a pivotal role in the control of COVID-19 pandemic. To carry out this strategy it is necessary to increase the testing capacity. Here, we compared a SARS CoV-2 rapid antigen test (RAT) and RT-PCR in 842 asymptomatic individuals from Tarapacá, Chile. We report a sensibility of 69.86%, a specificity of 99.61%, PPV of 94.44% and NPP of 97.22% with Ct values (Ct > 27) that were significantly higher among individuals with false-negative RAT. These results support the fact that RAT might have a significant impact in the identification of asymptomatic carriers in areas that lack well-equipped laboratories to perform SARS-CoV-2 real -time RT-PCR diagnostics or the results take more than 24-48 hours, as well as zones with high traffic of individuals, such as border/customs, airports, interregional bus, train stations or in any mass testing campaign requiring rapid results.

## Introduction

Given the increase in cases of SARS-CoV-2 infections worldwide, there is a need for a reliable rapid diagnostic test in addition to existing gold standard real-time RT-PCR. Rapid antigen tests (RAT) for SARS-CoV-2 can be performed onsite in mass testing, are inexpensive compared to real-time RT-PCR, do not require specific and expensive machinery and the results are available within 15 minutes (1), which could serve to evaluate chains of infection and their interruption. A meta-analysis recently revealed that the average sensitivity and specificity of the RAT for SARS-CoV-2 was 56.2% and 99.5%, respectively (2). Up to date, most of these validations were carried out in symptomatic individuals or samples previously collected (3-7). In contrast, onsite test validation studies in asymptomatic individuals are limited to supporting the use of RAT in mass testing and epidemiological surveillance (8-12).

### The Study

We evaluated the performance of the SARS-CoV-2 RAT (SD Biosensor, Inc. Republic of Korea, Catalog number 9901-NCOV-01G; Lateral flow immunoassay, Readout: visual-coloured bands, sensitivity of 96.52% and specificity of 99.68%) compared with the real-time RT-PCR for SARS-CoV-2 detection among asymptomatic individuals at Iquique city, Tarapacá Region, Chile. Samples were collected between January 14^th^ and January 17^th^, 2021, where the prevalence of SARS-CoV-2 infection was 11% in the region (13). Taking into consideration that the expected prevalence of positives for COVID-19 in asymptomatic individuals can vary from 8 to 12% and establishing a sampling error of 1.5% and a type I error of 5%, then the minimum size required was 864 ± 69,7. The sampling was carried out in seven testing sites corresponding to i) workers (n=56; 6.7%), ii) sanitary residence (n=239; 28.4%), and iii) general public (n=547; 65%). Overall, 854 individuals were included (mean age: 36.67 years, SD: 16.48 years; male n=51%; female n=41.6%; N/A n=7.4%). All participants completed a questionnaire and provided information on demographic characteristics, current and past (14 days) symptoms, and recent exposure to people with COVID-19 (Table 1). Two nasopharyngeal swabs (NSS) samples were collected by health care personnel at testing sites for each individual. NSS for RAT were analyzed according to the manufacturer’s instructions. NSS for real-time RT-PCR were analyzed within 24–72 hours of collection. Viral RNA was extracted using the Mag-Bind Viral DNA/RNA 96 kit (Omega Bio-Tek, Catalog number M6246) on the Kingfisher Flex Magnetic Particle Processor (Thermo Fisher Scientific). Real-time RT-PCR was performed using the GenomeCov19 Detection Kit ABM (Applied Biological Materials Inc, Canada, Catalog number G628.v2), with cycle threshold (Ct) values ≤40 considered positive for the N and S viral gene regions at Laboratorio Médico Bioclinic and Hospital Regional de Iquique. Statistical analysis considered sensitivity, specificity, Positive Predictive Value (PPV) and Negative Predictive Value (NPV), accuracy, and Kappa coefficient and Wilson score Confidence Interval at 95% (GraphPad Prism version 9.0.1).

**Table 1.**
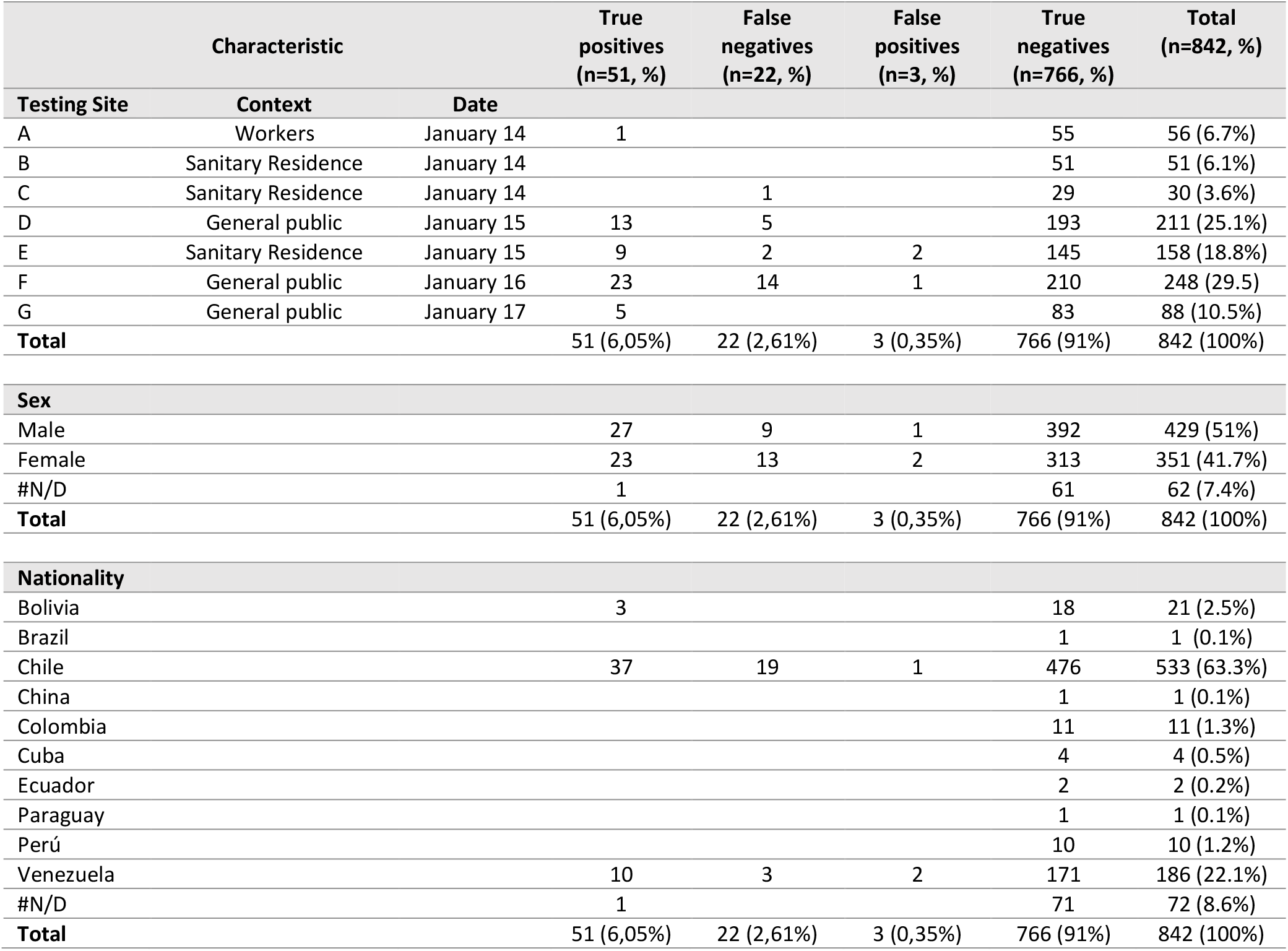
Characteristics of individuals providing paired NSS (N = 842) by results for SARS-CoV-2 real-time reverse transcription–polymerase chain reaction (RT-PCR) and Rapid Antigen Test (RAT) – January 2021. N/D: No Data

Among a total of 854 NSS submitted, 12 (1.4%) were excluded for lacking real-time RT-PCR results. Among 842 paired NSS from asymptomatic individuals, 54 (6.17%) were antigen-positive and 73 (8.6%) were real-time RT-PCR-positive. Antigen testing sensitivity was 69.86% (51 of 73), specificity was 99.61% (766 of 769), PPV was 94.44% (51 of 54), and NPV was 97.22% (766 of 789). Three paired (0.35%) NSS were antigen-positive and real-time RT-PCR– negative. Accuracy between the two techniques was 97.04% (Kappa coefficient = 0.78, 95% CI: 0.70 - 0.86). (Table 2). When we visualized the PCR Ct value data of the 73 samples that tested positive for NSS SARS-CoV-2 real-time RT-PCR detection, Ct values were significantly higher among individuals with false-negative RAT (viral gene N: 28.07 ± 4.343; viral gene S: 28.81 ± 3.873) compared to true positives (viral gene N: 19.99 ± 4.535; viral gene S: 20.93 ± 4.487) (Figure 1A). Next, we performed the k-means clustering analysis to find clusters in an iterative way using the Ct value of viral gene N (7). We identified three clusters: c1=16.87 (n=31, low Ct value), c2=24.5 (n=30, medium Ct value) and c3=31.67 (n=12, high Ct value), where 96.77% (30/31) of the samples with low Ct value also tested positive for RAT (Figure 1B, cluster 1) while the 66.6% (20/30) of the samples with medium Ct value also tested positive for RAT (Figure 1B, cluster 2). In contrast, only 8.33% (1/12) of the samples with high Ct value tested positive for RAT (Figure 1B, cluster 3). Only 3 RAT positive and SARS CoV-2 real-time RT-PCR negative samples were identified (Table 1).

**Table 2.**
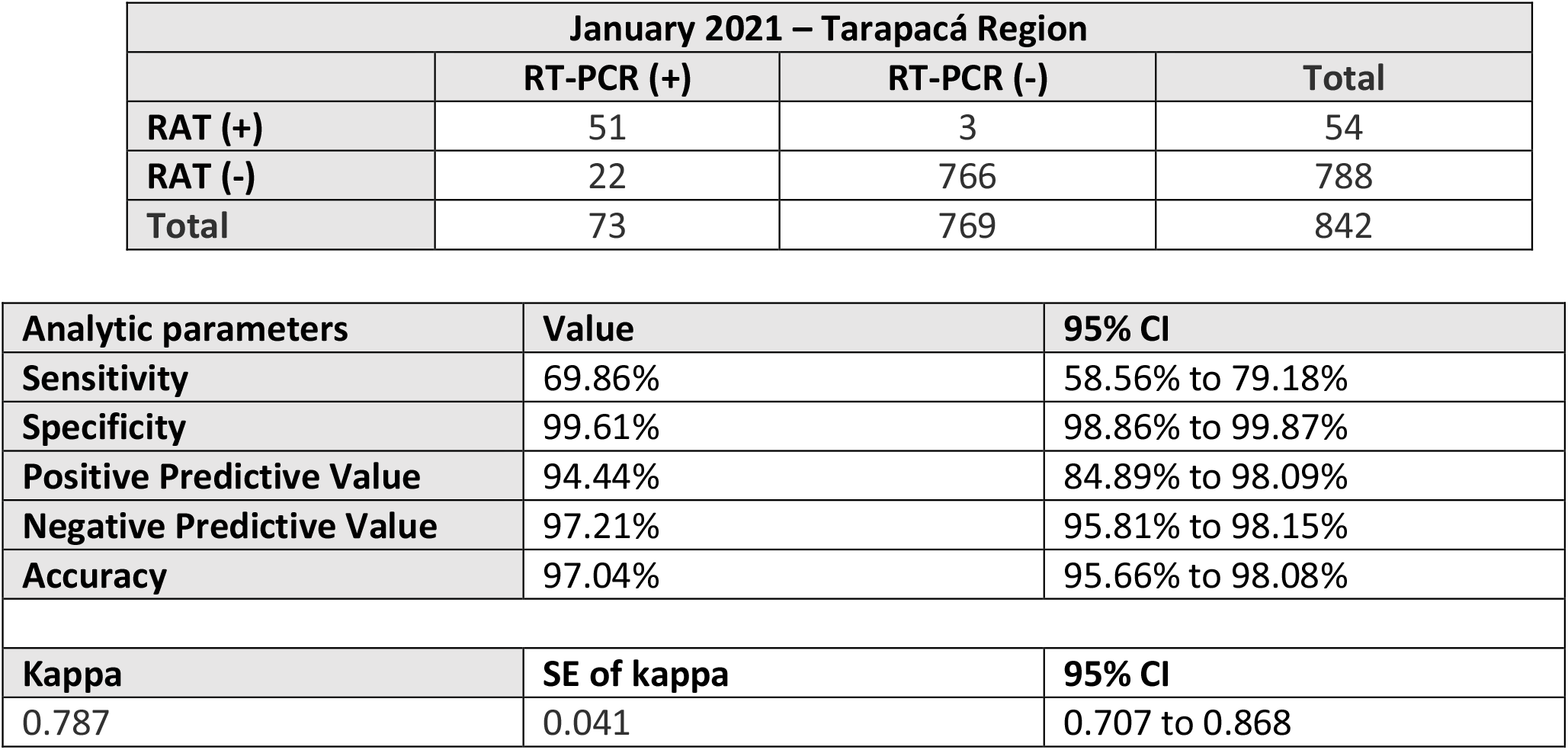
Agreement between RT-PCR test results and antigen test results overall.

**Figure 1:**
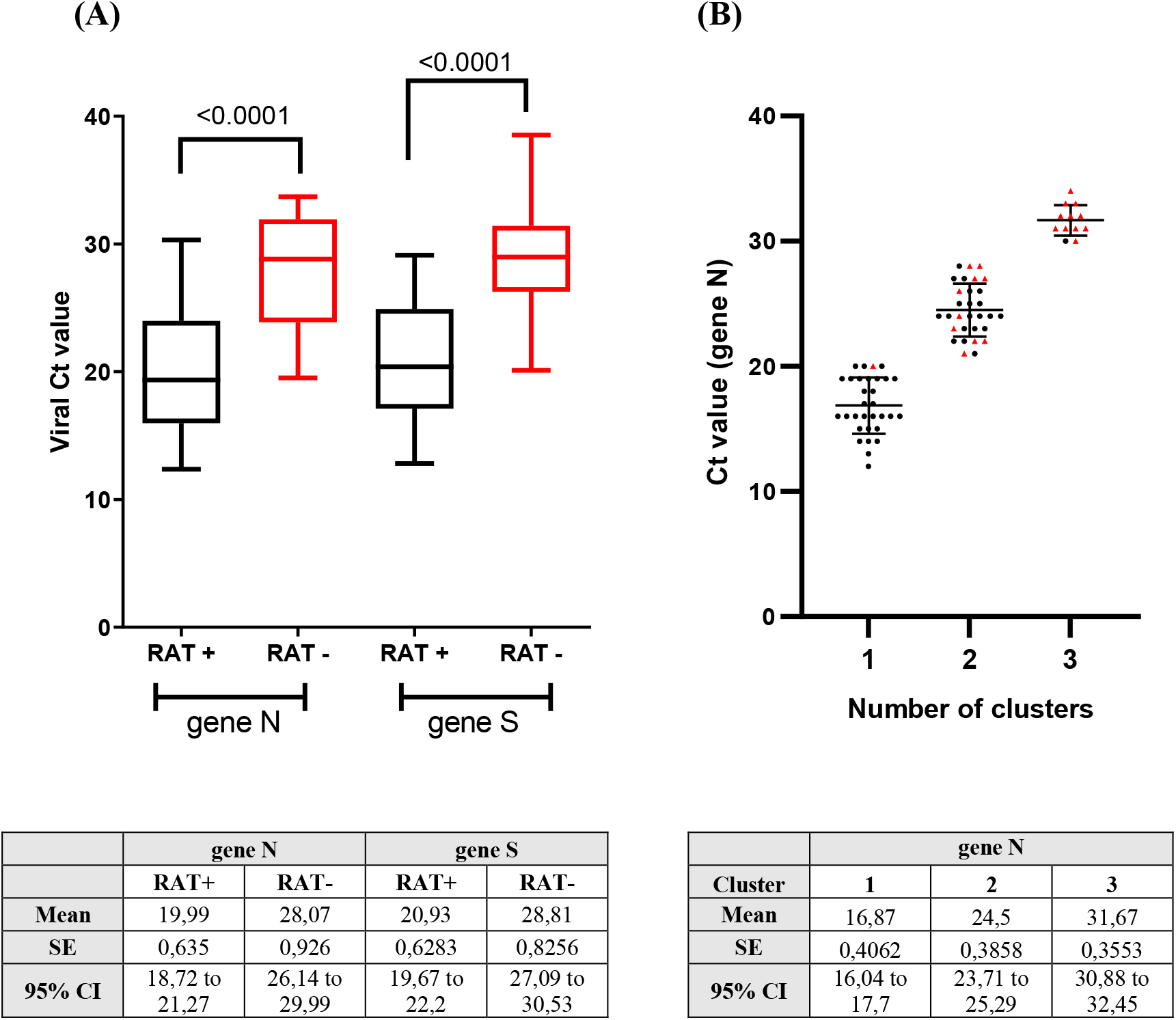
**(A)** Difference in viral cycle threshold (Ct) value between asymptomatic individuals with positive (black) and negative (red) RAT among real-time RT-PCR-positive (n=73). Analysis for statistical difference was performed by Wilcoxon test. (B) Plot of K-means clustering results of NSS real-time RT-PCR Ct values used to compare to RAT (black dots: RAT+ / red dots: RAT-).

## Conclusions

To control COVID-19 pandemic, improvement of SARS-CoV-2 diagnosis with easy, rapid, and cost-efficient approaches is urgently required. This study comprises asymptomatic individuals in three different contexts with an 8.64% prevalence of SARS-CoV-2 infection. In agreement with the Center of Disease Control and Prevention (CDC) recommendation on the use of antigen testing, the sensitivity of 69.86% indicates that RAT should not replace real-time RT-PCR in diagnosis and surveillance of SARS-CoV-2 infection (1). However, PPV of antigen testing was 94.44% indicating that asymptomatic persons with positive antigen results are infected with SARS-CoV-2 and would not require confirmatory real-time RT-PCR. Likewise, NPV of antigen testing was 97.21%, indicating that asymptomatic persons with negative antigen results are unlikely to be infected with SARS-CoV-2. Furthermore, individuals with false-negative results of the RAT had significantly higher Ct values (Ct > 27), which can be related to lower viral loads and less infectiousness in general (14, 15). Given the high predictive values in asymptomatic individuals and the fast test result implying faster tracing of infected individuals, these results support that RAT might have a significant role in COVID-19 screening, testing and contact tracing strategy to control COVID-19 pandemic in i) areas that lack well-equipped laboratories to perform SARS-CoV-2 real -time RT-PCR diagnostics, ii) areas where results take more than 24-48 hours, iii) areas with high traffic of individuals, such as border/customs, airports, interregional bus, train stations or in any mass testing campaign requiring rapid results.

## Data Availability

N/A

## Acknowledgments

We thank to Ministerio de Salud de Chile for funding this study. We gratefully acknowledge all the health services, hospitals and local health authorities that participated in study, Manuel Fernández (Seremi de Salud Tarapacá), Dolores Romero (Jefa de Salud Pública, Seremi de Salud), Jorge Galleguillos (Director Servicio de Salud Iquique), Carlos Calvo (Sub-Director Médico Servicio de Salud Iquique). Healthworkers from Departamento administración de Salud – Municipalidad de Iquique and Municipalidad de Alto Hospicio. The authors are supported by ANID Chile through Fondecyt grants N° 1181656 (A.G.), 1190156 (R.S-R.), 1180798 (F.V.-E.). F.V.-E., M.J.F., S.S., P.G., M.S.V., J.A., participated in the study design. M.P., M.A., C.G., performed the experiments. R.L., provided the NSS from COVID-19 asymptomatic individuals. F.P. performed the statistical analysis. F.V.-E., A.G., S.S., and R.S.-R. wrote the manuscript. M.S.V., and J.A., funding acquisition. All authors approved the final version of the manuscript.

## Ethical statement

Ministerio de Salud de Chile instructed this study to implement the use of the RAT in asymptomatic individuals. This work is considered thus for as a public health intervention to improve diagnosis. The study described here was approved by the Ethics Committee of the Faculty of Medicine at Universidad de Chile (Project N° 036-2020). NSS from COVID-19 positive and negative patients were anonymized.

## Notes

### Competing Interest Statement

The authors have declared no competing interest.

### Funding Statement

We thank to Ministerio de Salud de Chile for funding this study. The authors are supported by ANID Chile through Fondecyt grants N 1181656 (A.G.), 1190156 (R.S-R.), 1180798 (F.V.-E.).

### Author Declarations

Ministerio de Salud de Chile instructed this study to implement the use of the RAT in asymptomatic individuals. This work is considered thus for as a public health intervention to improve diagnosis. The study described here was approved by the Ethics Committee of the Faculty of Medicine at Universidad de Chile (Project N 036-2020). NSS from COVID-19 positive and negative patients were anonymized.

